# Biological Age Estimation Using Circulating Blood Biomarkers

**DOI:** 10.1101/2023.02.23.23285864

**Authors:** Jordan Bortz, Andrea Guariglia, Lucija Klaric, David Tang, Peter Ward, Michael Geer, Marc Chadeau-Hyam, Dragana Vuckovic, Peter K Joshi

## Abstract

Biological Age (BA) captures physiological deterioration better than chronological age and is amenable to interventions. Blood-based biomarkers have been identified as suitable candidates for BA estimation. This study aims to improve BA estimation using machine learning models and a feature-set of 60 circulating biomarkers available from the UK Biobank (UKBB) (n = 307,000). We implement an Elastic-Net derived Cox model with 25 selected biomarkers to predict mortality risk, which outperforms the well-known blood-biomarker based PhenoAge model, providing a 9.2% relative increase in predictive value. Importantly, we then show that using common clinical assay panels, with few biomarkers, alongside imputation and the model derived on the full set of biomarkers, does not substantially degrade predictive accuracy from the theoretical maximum achievable for the available biomarkers. BA is estimated as the equivalent age within the same-sex population which corresponds to an individual’s mortality risk. Values ranged between 20-years younger and 20-years older than individuals’ chronological age, exposing the magnitude of ageing signals contained in blood markers. Thus, we demonstrate a practical and cost-efficient method of estimating an improved measure of BA, available to the general population.

## MAIN

In the past few decades, BA has been estimated using a variety of biomarkers – telomere length, DNA-methylation, proteomics, metabolomics, glycomics, wearable sensor data and blood-based, clinical biomarkers (2, 3, 4, 5, 6, 7). Composite blood-based biomarkers have demonstrated an ability to detect differences in BA even in cohorts of young and healthy individuals, prior to the development of disease or phenotypic manifestations of accelerated ageing (8). When contrasted against some of the omics-based BA estimates, such as epigenetic clocks, blood biomarkers have considerable cost and scalability advantages (9, 10). However, the number of studies on blood biomarker-based BA estimation remains low and further validation is required (2, 11). By making use of a dataset of unprecedented size, this study aims to improve on BA estimation using machine learning methods and address real-world drawbacks, such as sparse data and cost.

Machine learning techniques have proved to be popular choices in the construction of BA estimates (2, 11, 12, 13). One of the most relevant studies in this domain is Levine *et al*.*’s* (1) 2018 paper, wherein the authors developed a BA measure called PhenoAge, using a Cox proportional-hazards model with an Elastic-Net penalty on data consisting of forty-two blood-based biomarkers and all-cause mortality collected within the NHANES programme. Liu *et al*. (14) subsequently demonstrated that PhenoAge was significantly associated with all-cause and cause-specific mortality, even after adjusting for chronological age (CA) and sex.

In this study, we used a dataset of 57 blood-biomarkers (Supplementary Table 1) and all-cause mortality from 306,756 participants from the UK Biobank (UKBB) dataset (https://www.ukbiobank.ac.uk/). Participants’ ages ranged from 37 to 73 years, with a mean of 56.3 years. The overall mortality rate of the population was 4.3% for females (6,515) and 7.8% for males (12,215), with a total of 18,730 recorded deaths. Follow-up duration ranged from 0.01 years to 14 years (average of 11.6 years) (Supplementary Table 2).

We built an Elastic-Net penalised Cox proportional-hazards model, as the foundation of our BA prediction, using survival time as outcome and blood biomarkers as predictors, while adjusting for sex and age, which were not penalised. As a non-linear alternative accounting for possible interactions we also constructed a Random Survival Forest (RSF) (15). Predictive performance of all models was evaluated using Harrell’s Concordance Index (“C-index” or “concordance statistic”), which is the most used measure of predictive discrimination in the context of survival models (16, 17).

We trained 100 Cox proportional-hazards models with Elastic-Net penalty, each on a 50% subsample of the training data (80-20 train-test split). Using a stability selection approach, we calculated the per-variable selection proportions across each of the 100 calibrated models as a proxy for their importance. We considered variables with selection proportion >80% to be stably selected (Fig 1a). A total of 25 (of the 57) biomarkers were stably selected, of which 22 had a selection proportion >95%. Reducing the selection proportion threshold to 50% would only have resulted in the inclusion of 4 additional variables. The ensemble of 25 stably selected features along with age and sex were included in a final unpenalized Cox model which we label the “Full Elastic-Net Cox (ENC)” model. The Full ENC model resulted in a C-Index of 0.762 with 95% confidence interval of [0.754 – 0.770] on the test set, showing improvement over (i) the model including sex and age only (null model) with a C-Index of 0.715 [0.707 – 0.723] and the PhenoAge model with C-Index of 0.739 [0.731 – 0.747] (Fig 1b).

**Fig 1:**
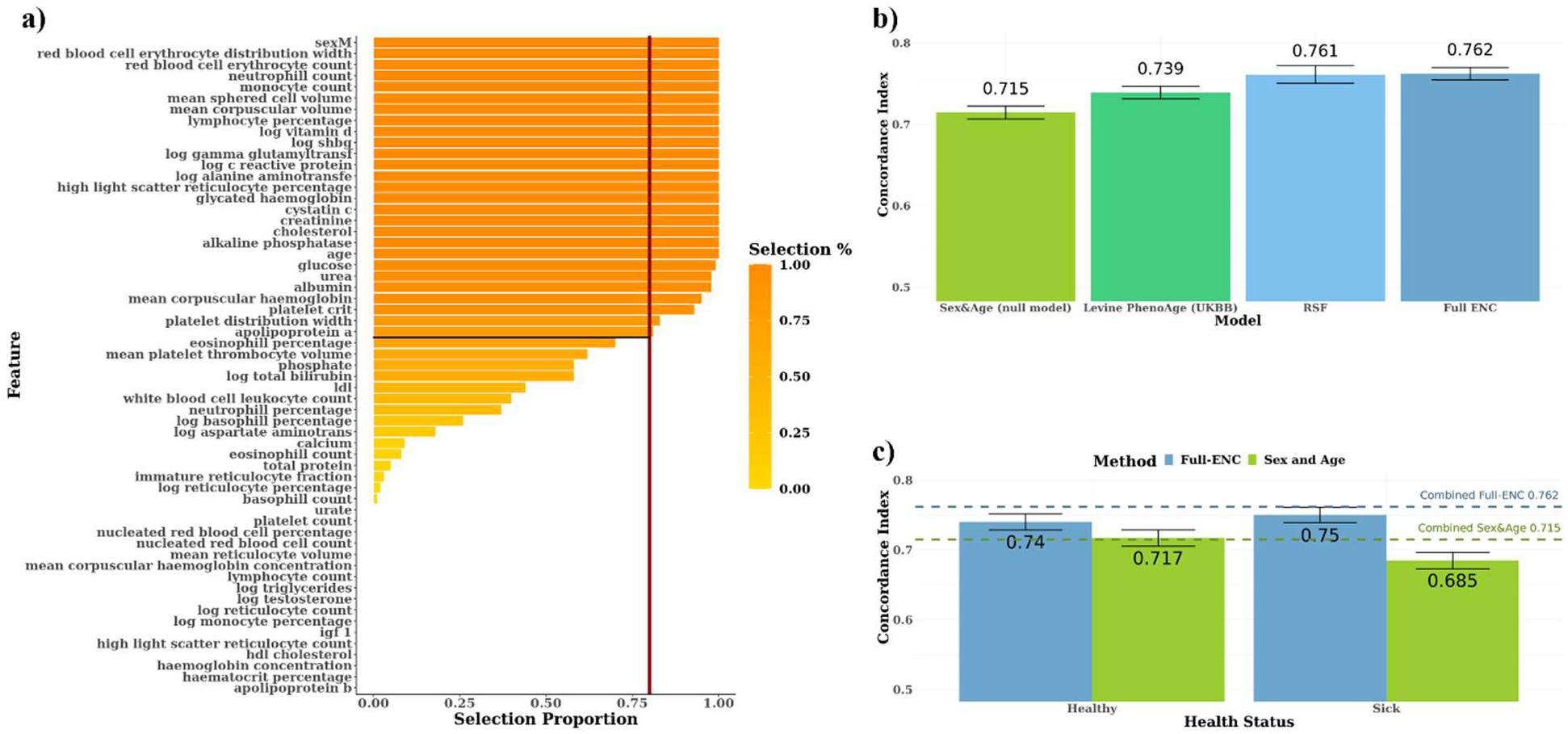
The Full Elastic-Net Cox model performs similarly to the Random Survival Forest and produces robust mortality risk predictions across both healthy and sick groups. (a) Selection proportions of each feature as a percentage of the 100 Elastic-Net iterations performed, ranked from highest to lowest. The adopted selection threshold of 80% is indicated in red. **(b)** Bar plot comparing the C-Index values (and 95% CI) of (1) a Cox model using sex and age only (null model), (2) the PhenoAge model applied on the UKBB data, (3) our RSF and (4) our Elastic-Net derived Cox-model (Full ENC). (**c)** Comparison of C-Index values of the Full ENC and sex-and-age-only null models for Healthy and Sick groups. For both groups, C-Index values of the Full ENC were significantly higher than those produced by the null model, with non-overlapping confidence intervals, indicating that the Full ENC model provides a statistically significant uplift in predictive ability. The dashed horizontal lines represent the C-index values of the Full ENC and the null model on the combined test set (Healthy + Sick).

The tuned RSF yielded a very similar result to the Full ENC model with a C-Index estimate of 0.761 [0.751 – 0.772] on the test set (Fig 1b), hence not supporting non-linear effects nor the existence of complex interactions across biomarkers. With a value of 0.5 corresponding to random prediction, we consider the predictive uplifts of models to be the additive increase above 0.5. The additive increases of 0.262 and 0.261 for the Full ENC and RSF models, compared to 0.215 for the null model, indicate a 22% and 21% increase in predictive value, respectively. Similarly, both models outperformed the PhenoAge model applied on the UKBB data, with a 0.023 increase in C-index (9.2% increase in predictive value).

As a sensitivity analysis to assess predictive ability and generalisability of the ENC model we stratified the test set by a self-reported, binary indicator of “long-standing illness, disability, or infirmity” (18, 19) and reported the corresponding C-Indices yielded by the null and ENC models trained on the full training dataset. The C-Index for those not reporting a prevalent condition at baseline (“Healthy”) was 0.740 [0.729 – 0.752] for the ENC model, which is higher than that from the null model at 0.717 [0.705 – 0.728]. In the group of participants reporting a prevalent condition (“Sick”) the estimated C-Index was comparable at 0.750 [0.739 – 0.761] for the ENC model and 0.685 [0.673 – 0.696] for the null model. In both groups of participants, the ENC model yielded an increase in the C-index compared to that of the null model, corresponding to 11% and 30% increases in predictive value, for the Healthy and Sick groups respectively. This stratified analysis suggests that the blood biomarkers detected by the ENC model are jointly predictive of mortality risk irrespective of prevalent morbidity.

Practical application of blood-based BA estimation will be facilitated by the use of existing results from assays which individuals have obtained for various clinical reasons. In principle, any subset of the 25 biomarkers might be available (i.e. >33 million combinations), although, in reality, clinical practice will tend to result in some subsets being more common than others. We researched which panels corresponded to the most performed blood tests in the UK as part of regular medical check-ups, employer-based testing, or diagnostic testing, within the NHS or by private providers (20, 21, 22) and identified the 10 most commonly available panels of markers (Supplementary Table 3). Similar, bespoke Elastic-Net Cox models were developed on the training set for each of these 10 representative panels of markers separately and we report the corresponding C-Indices in the test set (Fig 2, maroon bars). These C-Indices estimate the theoretical maximum of predictive accuracy for individuals who have had only these assays measured.

**Fig 2:**
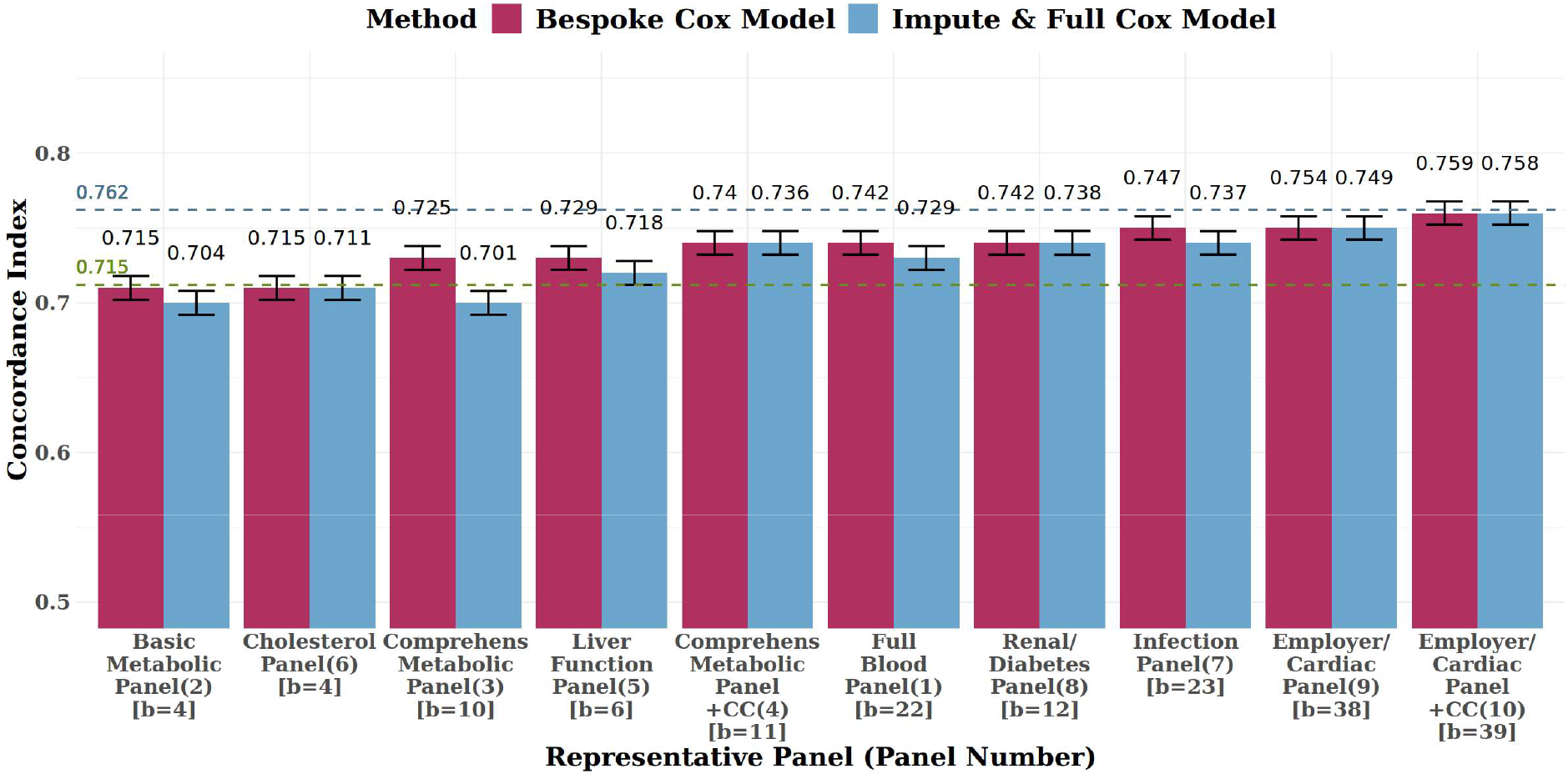
Imputing out-of-panel biomarkers and using the Full ENC model did not substantially reduce predictive accuracy compared to bespoke models for each representative panel. Comparison of concordance values across bespoke models vs imputed ENC models, for each of the 10 real-world representative blood panels, on the test set. Sex and age were also included as (unpenalised) features in all models. The number in brackets next to the panel name indicates the panel number as per Supplementary Table 3. The number in square brackets indicates the count of biomarkers (b=) in the panel. The performance of the impute-then-Full-ENC method is similar to that of the bespoke models, especially for the more comprehensive panels. The green dashed horizontal line indicates the 0.715 C-Index of the (sex and CA only) null model, whilst the blue line indicates the 0.762 C-Index of the Full ENC model with all 25 selected biomarkers measured. “+CC” indicates the addition of cystatin C to the panel (panels 4 and 10).

The performance of each of these per-panel models was compared to the performance of the Full ENC model, where we considered biomarkers not included in the panel to be unavailable, and were thus imputed using the k-Nearest Neighbour (kNN) method (23).

For all panels, except panel 3, there were no significant differences in C-Index values across both methods. The average reduction in C-index across all panels was 0.0087, whilst the reduction in C-index values for the top five performing panels (panels 1,8,7,9 and 10) was only 0.0036. These results appear to suggest that, for most panels, there is no substantial loss in predictive accuracy when imputing and using the ENC model compared to bespoke models.

After imputation, the ENC model for the top five performing panels yielded an average improvement in C-Index of 0.0273 over the sex-and-age-only null model (Fig 2, green line), and an average reduction in C-index of 0.0197 against the Full ENC model with all 25 selected biomarkers available (Fig 2, blue line). For panels 2 and 3, the ENC model yielded a C-index of 0.704 [0.696 – 0.712]and 0.701 [0.692 - 0.708], respectively, which is slightly lower than that of the sex-and-age-only model at 0.715 [0.707 – 0.723], suggesting that the imputation of missing variables was sufficiently imprecise to push predictions away from the point of utility.

Standardised model coefficients and 95-percent confidence intervals on the log hazard scale for the Full ENC model are displayed in Fig 3a. Increasing age, male sex and increased levels of cystatin C appear to have the strongest effects towards an increasing hazard rate, whilst high levels of creatinine, alanine aminotransferase and vitamin-D are associated with lower hazard rates. For the RSF, variable importance (VIMP) values were also generated. As was seen in the Full ENC model, age is the most important variable, followed by cystatin C, which appears to have a higher importance value than sex in the RSF (Supplementary Figure 2).

**Fig 3:**
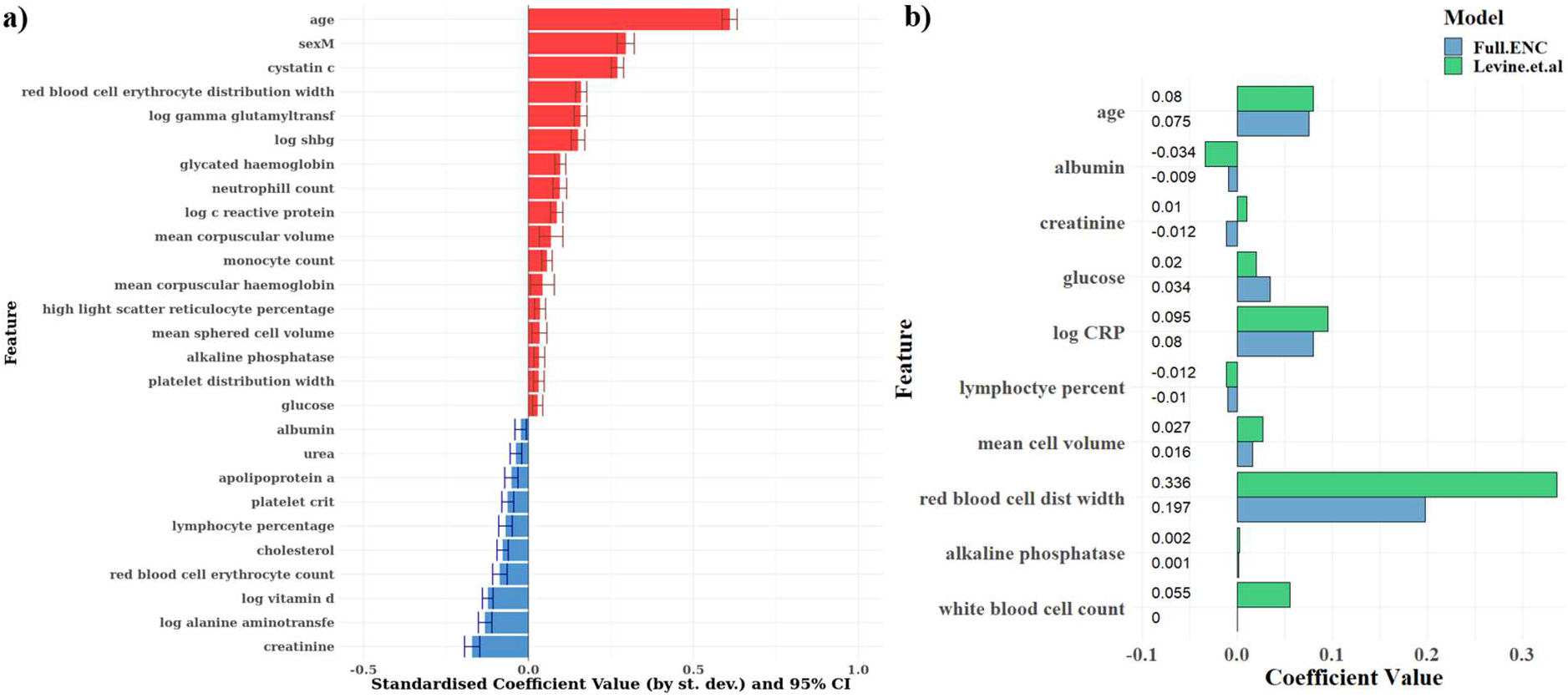
Our results confirm relationships suggested by Levine *et al*’s PhenoAge model, and additionally suggest that cystatin C is the biomarker of primary importance in BA estimation. **(a)** Bar chart showing standardised Cox model coefficients and 95% confidence intervals (log hazard scale) of the Full ENC model developed using stably selected variables, ranked in descending order. Coefficients are standardised (i.e. rescaled) by multiplying by the standard deviation of the variable concerned. Red indicates that higher levels increase mortality hazard; blue indicates that higher levels reduce mortality hazard. Apart from age and sex, cystatin C appears to have the strongest effect size. **(b)** Comparison of coefficient values between Levine *et. al’s* PhenoAge coefficients (green) and our Elastic-Net derived Cox model (blue). Model coefficients are similar across both models. Our ENC model selected individual WBC components (monocytes, neutrophils and lymphocytes) rather than overall WBC count. Measurement units for biomarkers were the same across both models.

The Full ENC model coefficients are compared to the coefficients as per Levine *et al*.’s PhenoAge (1) model in Fig 3b. The 10 variables in the PhenoAge model, derived from the NHANES dataset, were also stably selected in our ENC model except overall WBC count, which was substituted in our model by individual WBC components (monocytes, neutrophils, and lymphocytes). Model coefficients appear to be remarkably similar between the PhenoAge model and our ENC model, except creatinine, which had a small, negative effect size in our model, in contrast to a small, positive effect in the PhenoAge model. Overall, these consistent results provide validation of the effect that these variables in two large, independent cohorts. Our model additionally included sex and selected 17 additional biomarkers, some of which have the largest standardised coefficients and were not considered in the derivation of PhenoAge. The complete set of model coefficients is also shown in Supplementary Table 4 alongside implied hazard ratios.

The ENC model suggests that a one standard deviation increase (i) in age (8.1 years) increases mortality hazard by 84% [95% CI: 80% - 88%], (ii) in cystatin C (0.14mg/L) by 31% [29% - 33%], when adjusted for sex and all selected biomarkers (Supplementary Table 4). Our result mirrors the finding of a meta-analysis by Luo *et al*. (24), which identified a 32% [12% - 55%] increase in all-cause mortality hazard with a one standard deviation increase in cystatin C, based on 39,000 participants across nine different studies. The addition of cystatin C to the Comprehensive Metabolic Panel and the Employer/Cardiac Panel confirmed the incremental predictive uplifts in models where this biomarker is present, increasing the C-index by 0.015 and 0.005 for each of the Panels respectively (Fig 2, panels 3 vs 4 and 9 vs 10). Red blood cell (erythrocyte) distribution width appears to have the strongest effect size after cystatin C, with a standard deviation increase implying a 17% [15% – 19%] increase in mortality hazard.

Due to the documented positive relationship between serum cystatin C and creatinine (25, 26), as well as the positive correlation (r = 0.51) identified in our descriptive analysis (Supplementary Figure 1), we investigated how the effect sizes of these two biomarkers were altered with the removal of the other. When creatinine was removed, the standardised effect size for cystatin C was still the largest of all blood biomarkers. The hazard ratio decreased to a 22% [20% – 24%] increase in mortality hazard for one standard deviation increase in cystatin C level. Conversely, when cystatin C was removed, the effect size for creatinine was still negative but was no longer statistically significant, with a one standard deviation increase implying a 1% [-1% to 3%] reduction in mortality hazard.

The ENC model was used to estimate Biological Age Acceleration (BAA), representing the additional years of physiological deterioration above that implied by one’s CA (i.e., the difference between BA and CA), as explained in Methods. Most BAA values ranged from -20 to 20 years, with an interquartile range of 7 years, centered around a median value of –0.47 with a slight right skew (Fig 4a), similar to the results obtained by Liu *et al*. (14). However, the range of age acceleration seen in Liu *et al*. (14), of -3 to 6, is substantially narrower than our BAA range, which could at least partially be attributed to the differences in methodology used to derive BAA. It could also represent the additional sensitivity introduced in our model due to the inclusion of 17 additional circulating biomarkers, as well as the adjustment for sex.

**Fig 4:**
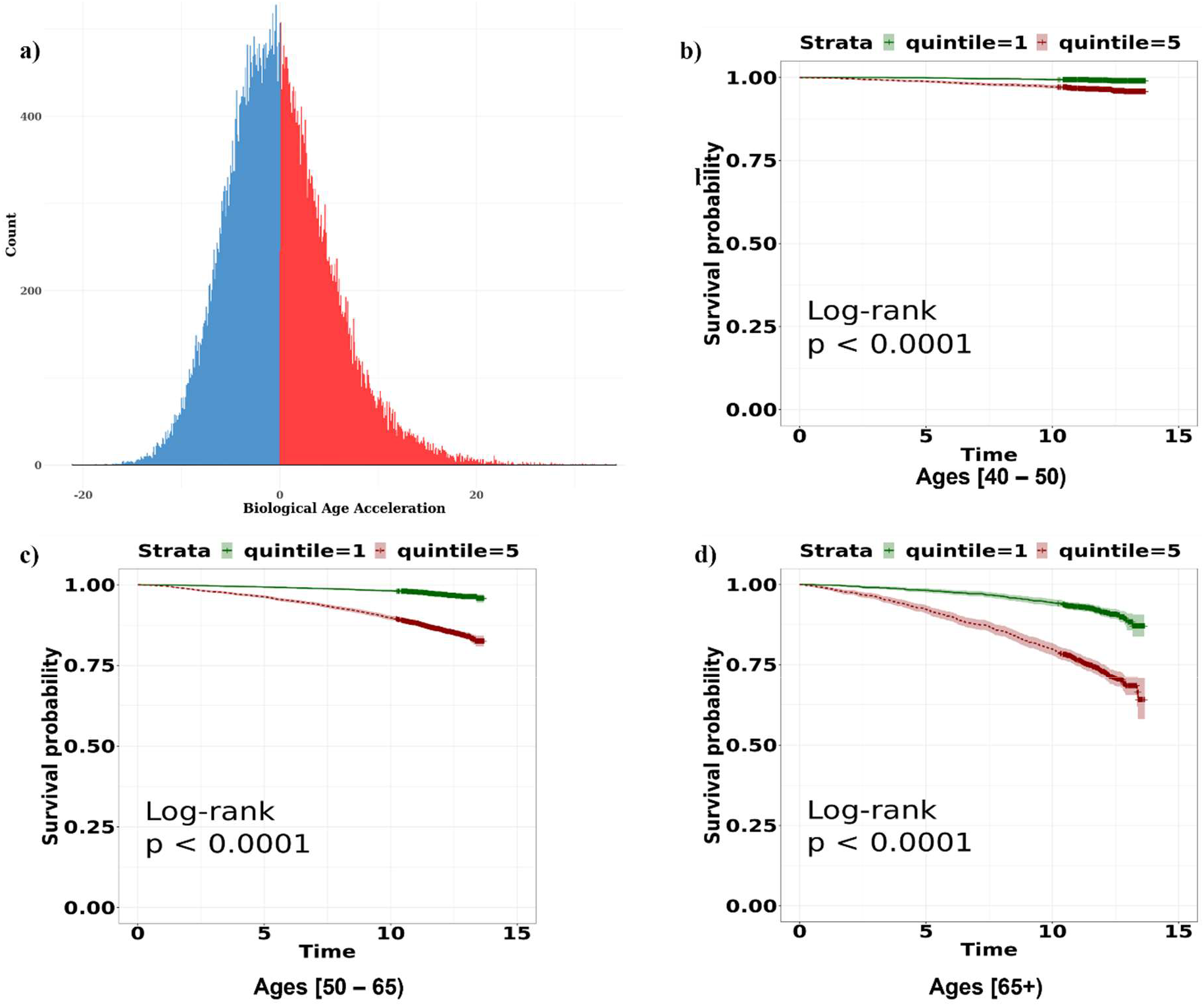
Biological Age Acceleration values range between -20 and 20, and reflect mortality risk even in same-age groups. **(a)** Distribution of estimated BAA values on the test set. Blue indicates a negative BAA (BA < CA), and red indicates a positive BAA (BA > CA). The distribution is largely symmetric around 0, with most values ranging between -20 and 20. (**b-d)** Kaplan-Meier curves comparing survival probabilities for the top and bottom BAA quintiles for each of the three age categories in the test set. Green indicates the bottom quintile (largest negative BAA values) whilst red indicates the top quintile (largest positive BAA values). Notable differences are observed between quintiles, especially at older ages.

As proposed by Liu *et al*. (14), we considered 3 broad age groups ([<=50], (50-65], (65+]), and plotted the Kaplan-Meier curves for the top and bottom quintiles of BAA values within each age group (Fig 4b-4d). The plots demonstrate that BAA provides an indication of BA and mortality risk even at the youngest ages in the dataset (<50 years of age at recruitment). The log-rank test was significant for all age groups (p < 0.0001). Notably, the mortality rates for those in the highest BAA quintile in an age group were higher than those in the lowest BAA quintile in the older age group. For example, those in the highest quintile of BAA in the <=50 age group experienced higher mortality rates than those in the lowest quintile of the 50 - 65 age group, over the next 14 years of follow-up.

As a final sensitivity analysis, we conducted similar analyses in males and females separately (Supplementary Note). The resulting C-index values, effect size estimates, and BAA predictions were similar to those obtained in the full population. Sex-stratified models did not produce more accurate predictions than the Full ENC model, even within same sex groups.

In this analysis, we demonstrated that circulating biomarkers have the potential to form the foundations of an accurate and low-cost measure of BA, via a simple formula. Our BA estimates ranged between 20-years younger and 20-years older than individuals’ chronological age, exposing the extent of ageing signals contained within the selected biomarkers. Our Elastic-Net derived Full ENC model, which selected 25 biomarkers, outperformed the alternative blood-biomarker based PhenoAge model (1), providing a 9.2% relative increase in predictive value. We do, however, acknowledge that our Full ENC model was both trained and evaluated within the UKBB cohort of participants.

Importantly, the model developed in this study possesses translational value in the real-world setting, where pre-existing measures of blood biochemistry will often be available but will vary greatly across individuals. We established that imputing values for unmeasured blood markers and subsequently using the Full ENC model, did not substantially degrade predictive accuracy away from what would be achieved by developing bespoke, per-panel models. The average reduction in C-Index across all panels, including the poor performing panels, was 0.0087, whilst the reduction for the top five panels was 0.0036. This suggests that the Full ENC model could be used to estimate BA values practically and accurately, in a real-world setting, irrespective of the number of biomarkers available. The ability of our BAA to distinguish between high risk and low risk individuals within same-age and same-health groups in our sample is an indication that BAA can identify physiological deterioration even in younger and healthy populations.

Our findings follow the trajectory of the growing literature in the field of BA, where numerous clocks have been developed which are predictive of future morbidity and mortality outcomes (2, 5). In particular, our focus on practicality and translational value was influenced by the blood-based PhenoAge (1) and GrimAge (27) BA estimators, which were shown to correlate with all-cause mortality even in same-age groups. We have demonstrated that such ageing clocks can indeed be employed highly cost effectively, using pre-existing or readily available blood results.

Our study benefited from several important strengths including the large sample size and the substantial number of biomarker measurements available, which enabled the development of robust inference, including stratified analyses and the use of interpretable machine learning techniques. However, the use of the UKBB dataset comes with well-documented limitations. The healthy volunteer bias inherent in the selection of participants, as well as the ethnic composition of participants (95% white), have been well documented in the literature (28). The homogeneity of the study population as well as the standardisation of measurement protocols across blood samples within the UKBB may not reflect systematic variation in blood results across different laboratories and populations. We do, however, acknowledge the remarkably similar parameters of Levine *et al’s* (1) PhenoAge model, which were based on the North-American, NHANES dataset, providing an independent validation of our findings using an external data source. An additional limitation relates to our imputation approach, which occurred within the single UKBB cohort. This approach may overestimate imputation quality in reality, where relationships between biomarkers in different population groups may differ from those observed in the UKBB.

Our findings are consequential in an era of rapidly ageing populations. Practical mechanisms for identifying biological ageing and scientifically informed ageing-reversal interventions are essential to maximising population healthspan and reducing pressure on healthcare providers.

## METHODS

### Data Source and Data Processing

The data used in this study forms part of the UK Biobank (UKBB) dataset. The 502,536 participants in the UKBB, recruited between 2006 and 2010 in England, Scotland, and Wales, were aged between 37 and 73 years at recruitment (29). Participants represented a range of socioeconomic, ethnic, and urban/rural population groups (29). Full details of the UKBB study, including details of recruitment and data collection, can be found on the UKBB website (https://www.ukbiobank.ac.uk/), and supplementary information can be obtained in Sudlow *et al*. (29). The specific data fields used in this analysis were date of recruitment, age at recruitment, sex, date of death (if applicable), presence of illness or disease at recruitment (self-reported), and a range of 60 blood biochemistry and haematology markers, samples of which were obtained from participants at recruitment. The full list of the blood biomarkers available in the UKBB, prior to the data cleaning and further sub-selection detailed below, can be found in Supplementary Table 1, alongside their measurement units. Additional details on these markers can be found in the UK Biobank Companion Documents (30, 31, 32).

Data fields were examined for missing values and any fields where more than 20% of observations contained missing values were removed (lipoprotein-A, oestradiol and rheumatoid factor). For the remaining fields, the underlying reasons for missingness were investigated, per biomarker, using the UKBB data showcase resources (https://biobank.ndph.ox.ac.uk/showcase/label.cgi?id=100080). Blood assay data in UKBB may be missing for a variety of reasons, such as aliquot or aliquot dilution problems, values being below the reportable limit, and no data being obtained from a sample. An examination of the underlying causes informed subsequent handling of data, and the appropriateness of proceeding under a complete case analysis. For example, if missing values were largely driven by measured levels being below the minimum limit of detection, omitting such individuals would exclude individuals with low levels of a biomarker from this study, biasing results. For all biomarkers, apart from testosterone, the reasons for missing values appeared to be independent of both the biomarker and the participant and were primarily caused by aliquot errors or aliquot dilution problems. We therefore proceeded under a complete case analysis, assuming that data was largely missing completely at random. The final dataset consisted of 306,756 observations, each having 57 blood biomarkers, alongside sex and age. Data was examined for outliers, and a log-transformation was applied to variables with strong positive skew (alanine aminotransferase, aspartate aminotransferase, gamma glutamyltransferase, C-reactive protein, sex hormone binding globulin (SHBG), testosterone, total bilirubin, triglyceride and vitamin-D). The final dataset was then randomly split into a training and test set in the ratio 80:20, and these were used in the remainder of the study.

### Model Performance

Predictive performance of all models was evaluated using Harrell’s Concordance Index (“C-index” or “concordance statistic”), which is the most commonly used measure of predictive discrimination in the context of survival models (16, 17). Concordance is defined as the probability that the magnitude of the predicted values for observations i and j (pi and pj), rank in the same order as the actual values for these observations, ai and aj:

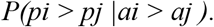

In other words, the probability that, for a pair of observations, the observation that experiences an event first, had a worse predicted outcome. The pair of observations is considered concordant if the rankings of the predicted and actual values are the same and are discordant otherwise. Ignoring ties, the Concordance Index is then defined as:

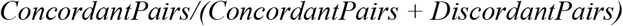

A concordance value of 0.5 corresponds to a model that is no better than a random guess, and a value of 1 would imply a perfect model. In the presence of censoring, some pairs cannot be compared or classified as concordant nor discordant. For example, pairing an individual who is censored at year 5 against an individual who experiences a mortality event at year 10 (17).

### Cox Proportional-Hazards Model with Elastic-Net Penalty and Stability Analysis

Using the training dataset, we developed 100 Elastic-Net Cox proportional-hazards models, using survival time and mortality indicator as outcome, each using a subsample of 50% of the dataset, as suggested by Bodinier *et al*. (33). For each iteration, 10-fold cross validation was used to select the optimal parameters, lambda and alpha, that minimised the partial likelihood deviance statistic, across a grid of 80 lambda values and alpha values between 0 and 1. As is commonly done in penalised regression models, we selected the (larger) lambda value that corresponded to a deviance statistic of one standard error above the minimum during cross-validation (lambda 1-s.e.) (34). The penalisation was only applied to the biomarker variables and neither sex nor age were penalised in any of the Elastic-Net models. All variables were standardised for the Elastic-Net optimisation process.

Stability analysis was performed to improve the predictive accuracy and the reliability of the model, ensuring that variable selection was not driven by outliers or a particular subset of observations (33). For each variable, we calculated the proportion of the 100 iterations in which the variable was selected, and its standardised coefficient value, as indicators of its relative importance. A selection proportion of 80% was used to determine if a variable was selected stably (35). These stably selected variables were then included in a non-penalised Cox model, to produce the final regression equation, which was used to generate predictions. This model is labelled the Full Elastic-Net Cox (ENC) model, where “full” indicates the use of all available biomarkers in model training along with age and sex.

This approach was used both for the model developed on the full set of biomarkers as well as for the 10 bespoke models developed for each of the 10 representative real-world blood panels. The Elastic-Net models were developed using the *survival* (36) and *glmnet* (37) packages in R version 4.1.3 (38).

### Biological Age Acceleration

We required an interpretable format of presenting an individual’s relative increase or decrease in mortality risk in relation to one’s CA, based on blood biochemistry. Biological Age Acceleration (BAA) is defined as the additive difference between one’s model-implied BA and one’s CA, adjusted for sex. To quantify this age acceleration, we decomposed the Cox model equation on the log scale as follows:

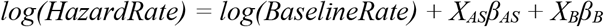

where X_AS_ represents the [n x 2] matrix of age and sex covariate values, β_AS_ represents the [2 × 1] vector of corresponding age and sex effect sizes (beta coefficients), X_B_ represents the [n x 57] matrix of biomarker values, β_B_ represents the [57 × 1] vector of corresponding biomarker effect sizes. Setting the first 2 terms on the righthand side of the equation as constant K, for an individual of a given age and sex:

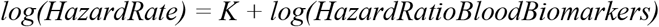

where *log(HazardRatioBloodBiomarkers)* is equivalent to the product X_*B*_β_*B*_. Using the relationship between log hazard ratio and age as suggested by Joshi *et al*. (39), where a log hazard ratio of Y indicates an additive change in age of approximately 10Y, we multiplied the product X_B_β_B_ by 10, to quantify the vector of BAA values. That is:

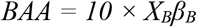

We independently checked this log-hazard-ratio:years-of-age relationship, by producing a Kaplan-Meier curve over age using the full UKBB dataset of 500,000 participants, and separately using data from the English Life Table No.17 (ELT-17) (40). Based on the results, we were satisfied that the relationship was sufficiently accurate to form the foundations of our BAA estimate. The results from this investigation can be seen in Supplementary Tables 5 – 7. Note that for the purposes of the BAA calculation, all blood biomarker variables in X_B_ were centred, by subtracting the (test set) mean value for each biomarker in the test set. This implies that BAA would equal to zero for a theoretical individual, of any age or sex, who holds the average value for each of the selected biomarkers.

Once BAA estimates had been derived as per the above method for the test set, we examined the range and distribution of BAA across observations. We also examined the association between BAA and mortality, by producing Kaplan-Meier curves for the lowest quintile and the highest quintile of BAA respectively. We did this across three age groups separately: 50 and younger, 50–65, and 65 years and older, which allowed for a comparison of mortality between BAA quintiles across different age groups. Kaplan-Meier curves were constructed using the *survival* (36) and *survminer* (41) packages in R.

### Random Survival Forest

A Random Survival Forest (RSF) was trained and the resulting C-Index value on the test set was compared to that achieved by the Full ENC model. A RSF is an extension of Breiman’s Random Forest method (42), applied to right-censored survival data (15). In the survival model setting, nodes are split according to the maximisation of the log-rank test statistic (15). The RSF has the potential to outperform the Cox model, as it can inherently handle nonlinear and interaction effects (15). We used the *randomForestSRC* (43) package in R to develop our RSF models.

Tuning was iteratively performed, maximising out-of-bag Concordance Index, for three hyper-parameters in particular: number of variables considered at each split, minimum node size and number of trees. The initial tuning was performed on a wide grid of these three hyper-parameter values, with each of the two successive rounds of tuning taking place over a smaller range of values, around the current optimal point. Three possible split points were considered for each continuous variable, at each split. The final optimal values for the number of variables considered at each split, and the minimum node size were 21 and 130 respectively, for an ensemble of 150 trees. The tuned model was then applied on the test set. The predicted outcome, per terminal node, is the Nelson-Aalen estimator for the cumulative hazard function, calculated on all the training observations in that terminal node. The average predicted cumulative hazard function value across all trees is then compared to the actual survival outcome, per pair of observations in the test set, to compute the C-Index.

We also produced Variable Importance (VIMP) scores, which provide an indication of which variables in the dataset hold the highest predictive ability (15). The VIMP ordering from the RSF was compared against the selection proportions and standardised coefficients from the Cox Elastic-Net stability analysis.

### Representative, Real World Blood Panels

We researched which panels corresponded to the most commonly performed blood tests in the UK as part of regular medical check-ups, employer-based testing, or diagnostic testing, within the NHS or by private providers (20, 21, 22). These panels are summarised in Supplementary Table 3. We decided to explore certain panels with and without the inclusion of cystatin C. This was done for two reasons. Firstly, cystatin C does not consistently form part of these blood panels; certain sources included it, whilst others did not. Secondly, during the course of our investigation, we observed the strong predictive ability of cystatin C in our own preliminary results. In light of these two reasons, we decided to test the sensitivity of performance between panels that did and did not include cystatin C.

We investigated the performance of the Full ENC model, applied on each of these representative panels. To do so, for each of the ten panels, we imputed the notionally unmeasured values of the biomarkers in the test set which were required by the Full ENC model, but were not present in the notional panel set, using k-Nearest Neighbours (kNN) imputation (k = 5) based on a random sample of 10,000 individuals from the training set. Variables were standardised for the KNN imputation, which was performed in R using the *VIM* package (23). For ease of reference, we label this approach the “impute-then-Full-ENC” method. In each of these 10 cases, the performance of the impute-then-Full-ENC prediction was compared against performance of each of 10 bespoke ENC models developed specifically on the biomarkers present in each of the ten representative panels, alongside sex and age. The bespoke models represent an estimate of the theoretical maximum of predictive accuracy for individuals who have had only these assays measured.

Note that a single, consistent test set was used to evaluate the performance of each and every one of the models in this study, removing any variability that would be introduced by altering the composition of the observations in the test set.

## Data Availability

The data used for this study is available to all qualified researchers via the UK Biobank data access process.

https://www.ukbiobank.ac.uk/

## CODE AVAILABILITY

The code used and resultant models for this project are the intellectual property of Humanity Inc.

## AUTHOR CONTRIBUTIONS

JB, PKJ, DV, DT, AG, PW, and MG participated in conceiving the study objectives and design. JB, PKJ and AG carried out initial data preparation. JB performed the data analysis and model development with support from PKJ, DV, LK, and DT. JB wrote the first draft of the manuscript. All authors revised and provided critical commentary on the manuscript. PKJ and DV jointly supervised this work.

## ACKOWNLEDGEMENTS

We thank the UK Biobank Resource, approved under application 69634. We acknowledge funding from Humanity Inc, a company dedicated to measuring and improving biological age. JB was supported by a Commonwealth Scholarship at Imperial College London, funded by the UK Foreign, Commonwealth & Development Office (FCDO). DV and MC-H are members of the Health Protection Research Unit in Chemical and Radiation Threats and Hazards, a partnership between Public Health England and Imperial College London which is funded by the National Institute for Health Research (NIHR). Neither the FCDO nor the NIHR had a role in the study design, data analysis, preparation of manuscript or decision to publish.

## ETHICS DECLARATIONS

PKJ and JB are paid consultants to Humanity Inc, a company focussed on measuring and developing interventions for Biological Age. LK is an employee of Humanity Inc. AG was formerly a paid consultant of Humanity Inc. MG and PW are founders of Humanity Inc and are employees and hold ordinary shares. PKJ, LK, MG and PW are partly remunerated under a Humanity Inc share option scheme. PKJ is founder of Geromica, a consultancy providing advice on measurement of health and aging. MC-H holds shares in the O-SMOSE company and has no conflict of interest to disclose. Consulting activities conducted by the company are independent of the present work.

Human subjects: This work used existing datasets, for which ethical approval had been gathered for health investigation at the time of collection.

## Notes

### Funding Statement

This study was partly funded by Humanity Inc, a company dedicated to measuring and improving biological age. JB was supported by a Commonwealth Scholarship at Imperial College London, funded by the UK Foreign, Commonwealth & Development Office (FCDO). DV and MC-H are members of the Health Protection Research Unit in Chemical and Radiation Threats and Hazards, a partnership between Public Health England and Imperial College London which is funded by the National Institute for Health Research (NIHR). Neither the FCDO nor the NIHR had a role in the study design, data analysis, preparation of manuscript or decision to publish.

### Author Declarations

UK Biobank has approval from the North West Multi-centre Research Ethics Committee (MREC) as a Research Tissue Bank (RTB) approval. This approval means that researchers do not require separate ethical clearance and can operate under the RTB approval. UK Biobank gave ethical approval for this work under application number 69634.

