## Supplementary Materials for "Biological Age Estimation Using Circulating Blood Biomarkers"

#### Supplementary Information

**Jordan Bortz (JB)[1,2,*]; Andrea Guariglia (AG)[1,2]; Lucija Klaric (LK)[1]; David Tang (DT)[2]; Peter Ward (PW)[1]; Michael Geer (MG)[1]; Marc Chadeau-Hyam (MC-H)[2,3,4]; Dragana Vuckovic (DV)[2,4,*,#]; Peter K Joshi (PKJ)[1,5,*,#]**

(1) Humanity Inc, Humanity, 177 Huntington Ave, Ste 1700, Humanity Inc - 91556, Boston, Massachusetts 02115, United States
(2) Department of Epidemiology and Biostatistics, School of Public Health, Faculty of Medicine, Imperial College London, London, UK
(3) MRC Centre for Environment and Health, School of Public Health, Imperial College London, London, UK
(4) NIHR-HPRU, Health Protection Research Unit in Chemical and Radiation Threats and Hazards, Public Health England and Imperial College London, UK
(5) Centre for Global Health Research, Usher Institute, University of Edinburgh, Edinburgh, UK

***Corresponding authors:**

**#Authors jointly supervised this work**

#### Supplementary Information

### Supplementary Tables

Supplementary Table 1: List of the 60 blood biomarkers available in the UK Biobank, split according to the UK Biobank definitions of Biochemistry and Haematology. Measurement units are displayed in adjacent columns. Highlighted in red are variables which exceeded the missingness threshold of 20% and were subsequently removed.

| | Blood Biochemistry (29) | Unit | Haematology (31) | Unit |
|---|---|---|---|---|
| 1 | Alanine Aminotransferase | U/L | Basophil number | x $10^9$ cells/L |
| 2 | Albumin | g/L | Basophil percent | % |
| 3 | Alkaline Phosphatase | U/L | Eosinophil number | x $10^9$ cells/L |
| 4 | Apolipoprotein A1 | g/L | Eosinophil percent | % |
| 5 | Apolipoprotein B | g/L | Haematocrit (relative volume of erythrocytes) | % |
| 6 | Aspartate Aminotransferase | U/L | Haemoglobin concentration | g/dL |
| 7 | Calcium | mmol/L | High Light scatter Reticulocytes % | % |
| 8 | Cholesterol | mmol/L | High Light scatter Reticulocytes count | x $10^{12}$ cells/L |
| 9 | Creatinine | µmol/L | Immature Reticulocyte Fraction | Ratio |
| 10 | Cystatin – C | mg/L | Lymphocyte number | x $10^9$ cells/L |
| 11 | Gamma-Glutamyltransferase | U/L | Lymphocyte percent | % |
| 12 | Glucose | mmol/L | Mean Corpuscular (erythrocyte) Haemoglobin | pg |
| 13 | Glycated haemoglobin | mmol/mol | Mean Corpuscular (erythrocyte) Hgb Conc | g/dL |
| 14 | High Density Lipoprotein (HDL) | mmol/L | Mean Corpuscular (erythrocyte) Volume | fL |
| 15 | High Sensitivity C-Reactive Protein (CRP) | mg/L | Mean Platelet (thrombocyte) Volume | fL |
| 16 | Insulin-like Growth Factor-1 (IGF-1) | nmol/L | Mean Reticulocyte Volume | fL |
| 17 | Lipoprotein a | nmol/L | Mean Sphered Cell Volume | fL |
| 18 | Low Density Lipoprotein (LDL) | mmol/L | Monocyte number | x $10^9$ cells/L |
| 19 | Oestradiol | pmol/L | Monocyte percent | % |
| 20 | Phosphate | mmol/L | Neutrophil number | x $10^9$ cells/L |
| 21 | Rheumatoid Factor | IU/ml | Neutrophil percent | % |
| 22 | Sex Hormone Binding Globulin | nmol/L | Nucleated Red Blood Cell number | x $10^9$ cells/L |
| 23 | Testosterone | nmol/L | Nucleated Red Blood Cell percent | % |
| 24 | Total Bilirubin | µmol/L | Platelet crit | % |
| 25 | Total Protein | g/L | Platelet Distribution Width | % |
| 26 | Triglyceride | mmol/L | Platelet or thrombocyte count | x $10^9$ cells/L |
| 27 | Urea | mmol/L | Red Blood Cell or erythrocyte count | x $10^{12}$ cells/L |
| 28 | Uric Acid | µmol/L | Red Cell (erythrocyte volume) Distribution Width | % |
| 29 | Vitamin D | nmol/L | Reticulocyte number | x $10^{12}$ cells/L |
| 30 | | | Reticulocyte percent | % |
| 31 | | | White Blood Cell or leukocyte count | x $10^9$ cells/L |

Supplementary Table 2: Descriptive statistics of non-blood variables, stratified by sex, and overall

| | Female (N=150 326) | Male (N=156 430) | Overall (N=306 756) |
|---|---|---|---|
| **Age** | | | |
| Mean (SD) | **55.8** (8.09) | **56.8** (8.19) | **56.3** (8.16) |
| Median [Min, Max] | **57.0** [39.0, 70.0] | **58.0** [37.0, 73.0] | **57.0** [37.0, 73.0] |
| **Death Event** | | | |
| No | **143 811** (95.7%) | **144 215** (92.2%) | **288 026** (93.9%) |
| Yes | **6 515** (4.3%) | **12 215** (7.8%) | **18 730** (6.1%) |
| **Survival/Follow-up Years** | | | |
| Mean (SD) | **11.7** (1.36) | **11.6** (1.71) | **11.6** (1.55) |
| Median [Min, Max] | **11.9** [0.0110, 14.0] | **11.8** [0.0137, 14.0] | **11.8** [0.0110, 14.0] |
| **Long Standing Illness/Disability** | | | |
| No | **106 026** (70.5%) | **100 538** (64.3%) | **206 564** (67.3%) |
| Yes | **40 210** (26.7%) | **52 300** (33.4%) | **92 510** (30.2%) |
| Do not know/ Prefer not to say | **4 090** (2.7%) | **3 592** (2.3%) | **7 682** (2.5%) |

Supplementary Table 3: Table showing the 10 representative blood panels, and which of the UK Biobank biomarkers are included in each. Some panels are comprised of markers included in other panels; these are indicated by the number in brackets.

| | Panel Name | Markers Included | | |
|---|---|---|---|---|
| 1 | **Full Blood Panel** | Basophil number<br>Basophil percent<br>Eosinophil number<br>Eosinophil percent<br>Haematocrit<br>Haemoglobin conc.<br>Lymphocyte number<br>Lymphocyte percent | Mean Corpuscular Haemoglobin<br>Mean Corpuscular Hgb Conc<br>Mean Corpuscular Volume<br>Mean Platelet Volume<br>Monocyte number<br>Monocyte percent<br>Neutrophil number | Neutrophil percent<br>Platelet crit<br>Platelet Distribution Width<br>Platelet or thrombocyte count<br>Red Blood Cell count<br>Red Cell Distribution Width<br>White Blood Cell |
| 2 | **Basic Metabolic Panel** | Glucose<br>Calcium | Urea<br>Creatinine | |
| 3 | **Comprehensive Metabolic Panel** | Glucose<br>Calcium<br>Albumin<br>Aspartate Aminotranferase | Urea<br>Creatinine<br>Total Protein | Alanine Aminotransferase<br>Total Bilirubin<br>Alkaline Phosphatase |
| 4 | **Comprehensive Metabolic Panel w Cystatin C** | As above, with Cystatin C | | |
| 5 | **Liver Function Panel** | Alanine Aminotransferase<br>Albumin | Aspartate Aminotransferase<br>Total Bilirubin | Alkaline Phosphatase<br>Gamma-Glutamyltransferase |
| 6 | **Cholesterol Panel** | Cholesterol<br>Triglyceride | High Density Lipoprotein (HDL) | Low Density Lipoprotein (LDL) |
| 7 | **Infection Panel** | Full Blood Panel (Panel 1) & High Sensitivity C-Reactive Protein (CRP) | | |
| 8 | **Renal Panel & Diabetic Testing** | Comprehensive Panel with Cystatin- C (Panel 4) & Glycated Haemoglobin | | |
| 9 | **Employer-based Test & Cardiac Panel** | Full Blood Panel (1)<br>Liver Function Panel (5) | Comprehensive Metabolic Panel wo Cystatin-C (3)<br>Cholesterol Panel (6) | <br>Renal Panel (8) |
| 10 | **Employer based Testing & Cardiac Panel w Cystatin C** | As above, with Cystatin-C | | |

Supplementary Table 4: Comparison of coefficient values between Levine *et al.*'s PhenoAge model and the Full ENC model. Levine *et al.*'s PhenoAge model (column A) included 10 variables, compared to 27 in our model (column B). Colours indicate the magnitude and direction of coefficients, with red indicating that higher levels increase mortality risk, whilst blue indicates a protective effect. For the variables selected by both models, coefficients are largely comparable, with the exception of creatinine. Measurement units of variables in both models are the same and can be found in Supplementary Table 1. The table also displays the standardised hazard ratios and 95% CIs of stably selected variables as per the Full ENC model, in columns C and D.

| Feature | Levine et. al. (A) | Full ENC model coefficients (B) | Hazard Ratio per SD increase (C) | Hazard Ratio 95% CI (D) |
|---|---|---|---|---|
| Age | 0.08 | 0.075 | 1.84 | 1.80 - 1.88 |
| Albumin | -0.034 | -0.009 | 0.98 | 0.96 - 0.99 |
| Creatinine | 0.0095 | -0.0117 | 0.84 | 0.82 - 0.86 |
| Glucose | 0.0195 | 0.0344 | 1.03 | 1.01 - 1.04 |
| log CRP | 0.0954 | 0.08 | 1.09 | 1.07 - 1.11 |
| Lymphocyte % | -0.012 | -0.010 | 0.93 | 0.91 - 0.95 |
| Mean cell vol | 0.0268 | 0.0162 | 1.07 | 1.03 - 1.11 |
| Red Cell Distribution Width | 0.3356 | 0.1973 | 1.17 | 1.15 - 1.19 |
| Alkaline Phosphatase | 0.00188 | 0.00147 | 1.03 | 1.02 - 1.05 |
| White Blood Cell Count | 0.0554 | <50% selection | | |
| Apolipoprotein A | | -0.198 | 0.95 | 0.93 - 0.97 |
| Cholesterol | | -0.078 | 0.92 | 0.91 - 0.94 |
| Cystatin-C | | 1.877 | 1.31 | 1.29 - 1.33 |
| Glycated Haemoglobin | | 0.019 | 1.10 | 1.08 - 1.12 |
| High Light Scatter Reticulocyte % | | 0.178 | 1.04 | 1.02 - 1.05 |
| log Vitamin D | | -0.260 | 0.88 | 0.87 - 0.9 |
| log Alanine Aminotransferase | | -0.300 | 0.88 | 0.86 - 0.9 |
| log Gamma-Glutamyltransferase | | 0.264 | 1.17 | 1.15 - 1.19 |
| log Sex Hormone Binding Globulin | | 0.310 | 1.16 | 1.14 - 1.19 |
| Mean Corpuscular Haemoglobin | | 0.026 | 1.04 | 1.01 - 1.08 |
| Mean Sphered Cell Volume | | 0.007 | 1.03 | 1.01 - 1.06 |
| Monocyte Count | | 0.340 | 1.06 | 1.04 - 1.07 |
| Neutrophill Count | | 0.070 | 1.10 | 1.08 - 1.12 |
| Platelet Crit | | -1.356 | 0.94 | 0.92 - 0.96 |
| Platelet Distribution Width | | 0.060 | 1.03 | 1.02 - 1.05 |
| Red Blood Cell Count | | -0.215 | 0.92 | 0.90 - 0.94 |
| Sex (male) | | 0.590 | 1.34 | 1.31 - 1.38 |
| Urea | | -0.030 | 0.96 | 0.95 - 0.98 |

Supplementary Table 5: Life table based on Kaplan-Meier curve for entire UKBB dataset, with an illustration of the relationship between log hazard ratio and age as suggested by Joshi *et al.* (39), where a log hazard ratio of Y indicates an additive change in age of approximately 10Y, for intervals of 2, 5 and 7 years. Ages 50 – 75 shown. The row above age 50 illustrates the calculation dynamics for Age 50 (first row of main table). Bottom right shows the average of the log hazard ratio columns, and should equate to 0.2, 0.5 and 0.7 respectively.

**UKBB mortality table based on Kaplan-Meier curve.**
**Males and females combined, ages 50 - 75**

| Age | $_{n}p_{50}$ | $q_x$ | $m_x$ | $m_x$ ratio:2yr | $m_x$ ratio:5yr | $m_x$ ratio:7yr | LN($HR_2$) | LN($HR_5$) | LN($HR_7$) |
|---|---|---|---|---|---|---|---|---|---|
| A | B | C | D | E | F | G | H | I | J |
| *Calc Age 50 (row1):* | From KM curve | 1 - $B_{51}/B_{50}$ | -LN(1-C) | $D_{52}/D_{50}$ | $D_{55}/D_{50}$ | $D_{57}/D_{50}$ | LN( E ) | LN( F ) | LN( G ) |
| **50** | 0.991 | 0.001 | 0.001 | 1.406 | 1.563 | 1.885 | 0.34 | 0.45 | 0.63 |
| **51** | 0.989 | 0.001 | 0.001 | 1.113 | 1.478 | 1.933 | 0.11 | 0.39 | 0.66 |
| **52** | 0.988 | 0.002 | 0.002 | 1.083 | 1.341 | 1.561 | 0.08 | 0.29 | 0.45 |
| **53** | 0.986 | 0.002 | 0.002 | 1.264 | 1.737 | 1.850 | 0.23 | 0.55 | 0.62 |
| **54** | 0.985 | 0.002 | 0.002 | 1.079 | 1.440 | 1.696 | 0.08 | 0.36 | 0.53 |
| **55** | 0.983 | 0.002 | 0.002 | 1.206 | 1.464 | 1.709 | 0.19 | 0.38 | 0.54 |
| **56** | 0.981 | 0.002 | 0.002 | 1.308 | 1.572 | 1.970 | 0.27 | 0.45 | 0.68 |
| **57** | 0.979 | 0.002 | 0.002 | 1.164 | 1.417 | 1.753 | 0.15 | 0.35 | 0.56 |
| **58** | 0.976 | 0.003 | 0.003 | 1.065 | 1.506 | 1.737 | 0.06 | 0.41 | 0.55 |
| **59** | 0.973 | 0.003 | 0.003 | 1.177 | 1.506 | 1.869 | 0.16 | 0.41 | 0.63 |
| **60** | 0.971 | 0.003 | 0.003 | 1.167 | 1.631 | 1.977 | 0.15 | 0.49 | 0.68 |
| **61** | 0.968 | 0.003 | 0.003 | 1.253 | 1.588 | 1.958 | 0.23 | 0.46 | 0.67 |
| **62** | 0.964 | 0.004 | 0.004 | 1.237 | 1.694 | 2.100 | 0.21 | 0.53 | 0.74 |
| **63** | 0.961 | 0.004 | 0.004 | 1.153 | 1.562 | 1.955 | 0.14 | 0.45 | 0.67 |
| **64** | 0.957 | 0.004 | 0.004 | 1.241 | 1.698 | 2.175 | 0.22 | 0.53 | 0.78 |
| **65** | 0.953 | 0.005 | 0.005 | 1.212 | 1.695 | 2.189 | 0.19 | 0.53 | 0.78 |
| **66** | 0.948 | 0.005 | 0.005 | 1.233 | 1.752 | 2.219 | 0.21 | 0.56 | 0.8 |
| **67** | 0.943 | 0.006 | 0.006 | 1.240 | 1.805 | 2.205 | 0.21 | 0.59 | 0.79 |
| **68** | 0.937 | 0.007 | 0.007 | 1.251 | 1.800 | 2.258 | 0.22 | 0.59 | 0.81 |
| **69** | 0.931 | 0.007 | 0.007 | 1.281 | 1.779 | 2.278 | 0.25 | 0.58 | 0.82 |
| **70** | 0.924 | 0.008 | 0.008 | 1.291 | 1.805 | 2.255 | 0.26 | 0.59 | 0.81 |
| **71** | 0.917 | 0.009 | 0.009 | 1.266 | 1.778 | 2.020 | 0.24 | 0.58 | 0.7 |
| **72** | 0.908 | 0.011 | 0.011 | 1.221 | 1.746 | 2.014 | 0.2 | 0.56 | 0.7 |
| **73** | 0.898 | 0.012 | 0.012 | 1.254 | 1.595 | 1.981 | 0.23 | 0.47 | 0.68 |
| **74** | 0.888 | 0.013 | 0.013 | 1.281 | 1.649 | 1.698 | 0.25 | 0.5 | 0.53 |
| **75** | 0.876 | 0.015 | 0.015 | 1.249 | 1.579 | 0.955 | 0.22 | 0.46 | -0.05 |
| | | | | | | **Average LN(Hazard Ratio):** | **0.20** | **0.48** | **0.67** |
| | | | | | | **Joshi et al.'s suggestion:** | **0.20** | **0.50** | **0.70** |

Supplementary Table 6: As above, for ELT-17 life table; males only. Ages 50 – 75 shown. The row above age 50 illustrates the calculation dynamics for Age 50 (first row of main table). Bottom right shows the average of the log hazard ratio columns, and should equate to 0.2, 0.5 and 0.7 respectively.

**English Life Tables No 17; period expectation of life.**
**Based on data for England and Wales for the years 2010-2012**
**Males**

| Age | $m_x$ | $m_x$ ratio:2yr | $m_x$ ratio:5yr | $m_x$ ratio:7yr | LN($HR_2$) | LN($HR_5$) | LN($HR_7$) |
|---|---|---|---|---|---|---|---|
| A | B | C | D | E | F | G | H |
| *Calc Age 50 (row1):* | From ELT 17 | $C_{52}/C_{50}$ | $C_{55}/C_{50}$ | $C_{57}/C_{50}$ | LN( C ) | LN( D ) | LN( E ) |
| **50** | 0.003 | 1.221 | 1.627 | 1.978 | 0.20 | 0.49 | 0.68 |
| **51** | 0.003 | 1.216 | 1.630 | 1.953 | 0.20 | 0.49 | 0.67 |
| **52** | 0.004 | 1.208 | 1.620 | 1.929 | 0.19 | 0.48 | 0.66 |
| **53** | 0.004 | 1.211 | 1.606 | 1.927 | 0.19 | 0.47 | 0.66 |
| **54** | 0.005 | 1.221 | 1.597 | 1.921 | 0.20 | 0.47 | 0.65 |
| **55** | 0.005 | 1.216 | 1.590 | 1.885 | 0.20 | 0.46 | 0.63 |
| **56** | 0.006 | 1.198 | 1.574 | 1.842 | 0.18 | 0.45 | 0.61 |
| **57** | 0.006 | 1.191 | 1.550 | 1.835 | 0.17 | 0.44 | 0.61 |
| **58** | 0.007 | 1.200 | 1.537 | 1.860 | 0.18 | 0.43 | 0.62 |
| **59** | 0.007 | 1.203 | 1.541 | 1.879 | 0.18 | 0.43 | 0.63 |
| **60** | 0.008 | 1.185 | 1.551 | 1.879 | 0.17 | 0.44 | 0.63 |
| **61** | 0.009 | 1.170 | 1.562 | 1.899 | 0.16 | 0.45 | 0.64 |
| **62** | 0.010 | 1.184 | 1.585 | 1.962 | 0.17 | 0.46 | 0.67 |
| **63** | 0.010 | 1.210 | 1.623 | 2.028 | 0.19 | 0.48 | 0.71 |
| **64** | 0.011 | 1.219 | 1.657 | 2.048 | 0.20 | 0.51 | 0.72 |
| **65** | 0.012 | 1.212 | 1.675 | 2.035 | 0.19 | 0.52 | 0.71 |
| **66** | 0.014 | 1.216 | 1.680 | 2.025 | 0.20 | 0.52 | 0.71 |
| **67** | 0.015 | 1.237 | 1.680 | 2.027 | 0.21 | 0.52 | 0.71 |
| **68** | 0.017 | 1.249 | 1.665 | 2.028 | 0.22 | 0.51 | 0.71 |
| **69** | 0.019 | 1.236 | 1.638 | 2.018 | 0.21 | 0.49 | 0.70 |
| **70** | 0.021 | 1.215 | 1.623 | 2.012 | 0.19 | 0.48 | 0.70 |
| **71** | 0.023 | 1.205 | 1.633 | 2.027 | 0.19 | 0.49 | 0.71 |
| **72** | 0.025 | 1.207 | 1.656 | 2.066 | 0.19 | 0.50 | 0.73 |
| **73** | 0.028 | 1.218 | 1.682 | 2.122 | 0.20 | 0.52 | 0.75 |
| **74** | 0.031 | 1.231 | 1.711 | 2.177 | 0.21 | 0.54 | 0.78 |
| **75** | 0.034 | 1.240 | 1.742 | 2.214 | 0.21 | 0.56 | 0.79 |
| | | **Average LN(Hazard Ratio):** | | | **0.19** | **0.48** | **0.68** |
| | | **Joshi et al.'s suggestion:** | | | **0.20** | **0.50** | **0.70** |

Supplementary Table 7: ELT-17 life table as above; females only

**Females**

| x | $m_x$ | $m_x$ ratio:2yr | $m_x$ ratio:5yr | $m_x$ ratio:7yr | LN($HR_2$) | LN($HR_5$) | LN($HR_7$) |
|---|---|---|---|---|---|---|---|
| **A** | **B** | **C** | **D** | **E** | **F** | **G** | **H** |
| *Calc Age 50 (row1):* | **From ELT 17** | $C_{52}/C_{50}$ | $C_{55}/C_{50}$ | $C_{57}/C_{50}$ | **LN( C )** | **LN( D )** | **LN( E )** |
| **50** | 0.002 | 1.218 | 1.580 | 1.895 | 0.20 | 0.46 | 0.64 |
| **51** | 0.002 | 1.202 | 1.559 | 1.882 | 0.18 | 0.44 | 0.63 |
| **52** | 0.003 | 1.190 | 1.556 | 1.878 | 0.17 | 0.44 | 0.63 |
| **53** | 0.003 | 1.187 | 1.566 | 1.876 | 0.17 | 0.45 | 0.63 |
| **54** | 0.003 | 1.191 | 1.579 | 1.861 | 0.17 | 0.46 | 0.62 |
| **55** | 0.003 | 1.200 | 1.581 | 1.834 | 0.18 | 0.46 | 0.61 |
| **56** | 0.004 | 1.207 | 1.562 | 1.813 | 0.19 | 0.45 | 0.60 |
| **57** | 0.004 | 1.207 | 1.529 | 1.806 | 0.19 | 0.42 | 0.59 |
| **58** | 0.004 | 1.198 | 1.502 | 1.805 | 0.18 | 0.41 | 0.59 |
| **59** | 0.005 | 1.179 | 1.495 | 1.809 | 0.16 | 0.40 | 0.59 |
| **60** | 0.005 | 1.160 | 1.507 | 1.828 | 0.15 | 0.41 | 0.60 |
| **61** | 0.006 | 1.160 | 1.534 | 1.877 | 0.15 | 0.43 | 0.63 |
| **62** | 0.006 | 1.181 | 1.576 | 1.945 | 0.17 | 0.45 | 0.67 |
| **63** | 0.007 | 1.202 | 1.617 | 2.001 | 0.18 | 0.48 | 0.69 |
| **64** | 0.007 | 1.210 | 1.647 | 2.027 | 0.19 | 0.50 | 0.71 |
| **65** | 0.008 | 1.213 | 1.665 | 2.037 | 0.19 | 0.51 | 0.71 |
| **66** | 0.009 | 1.223 | 1.676 | 2.047 | 0.20 | 0.52 | 0.72 |
| **67** | 0.010 | 1.235 | 1.679 | 2.062 | 0.21 | 0.52 | 0.72 |
| **68** | 0.011 | 1.237 | 1.674 | 2.081 | 0.21 | 0.51 | 0.73 |
| **69** | 0.012 | 1.231 | 1.670 | 2.106 | 0.21 | 0.51 | 0.74 |
| **70** | 0.013 | 1.224 | 1.682 | 2.142 | 0.20 | 0.52 | 0.76 |
| **71** | 0.015 | 1.222 | 1.711 | 2.187 | 0.20 | 0.54 | 0.78 |
| **72** | 0.016 | 1.228 | 1.750 | 2.239 | 0.21 | 0.56 | 0.81 |
| **73** | 0.018 | 1.243 | 1.791 | 2.292 | 0.22 | 0.58 | 0.83 |
| **74** | 0.020 | 1.261 | 1.824 | 2.343 | 0.23 | 0.60 | 0.85 |
| **75** | 0.023 | 1.273 | 1.844 | 2.386 | 0.24 | 0.61 | 0.87 |
| | | | **Average LN(Hazard Ratio):** | | **0.19** | **0.49** | **0.69** |
| | | | **Joshi et al.'s suggestion:** | | **0.20** | **0.50** | **0.70** |

### Supplementary Figures

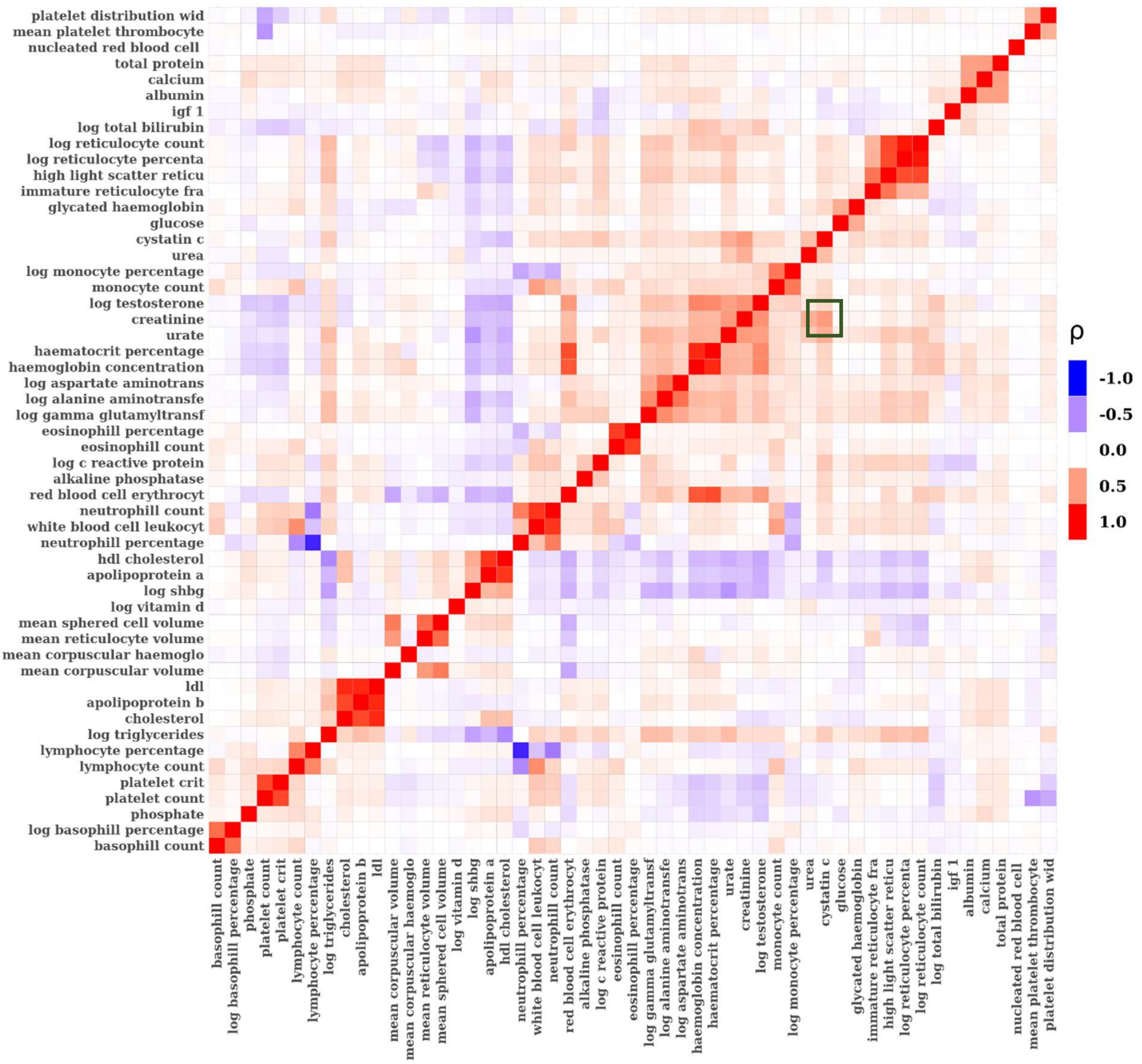


Fig SF1: Correlation heatmap of all blood biomarkers. Red indicates a strong positive correlation; blue indicates a strong negative correlation; white indicates no correlation. We note the positive correlation between creatinine and cystatin C (green square).

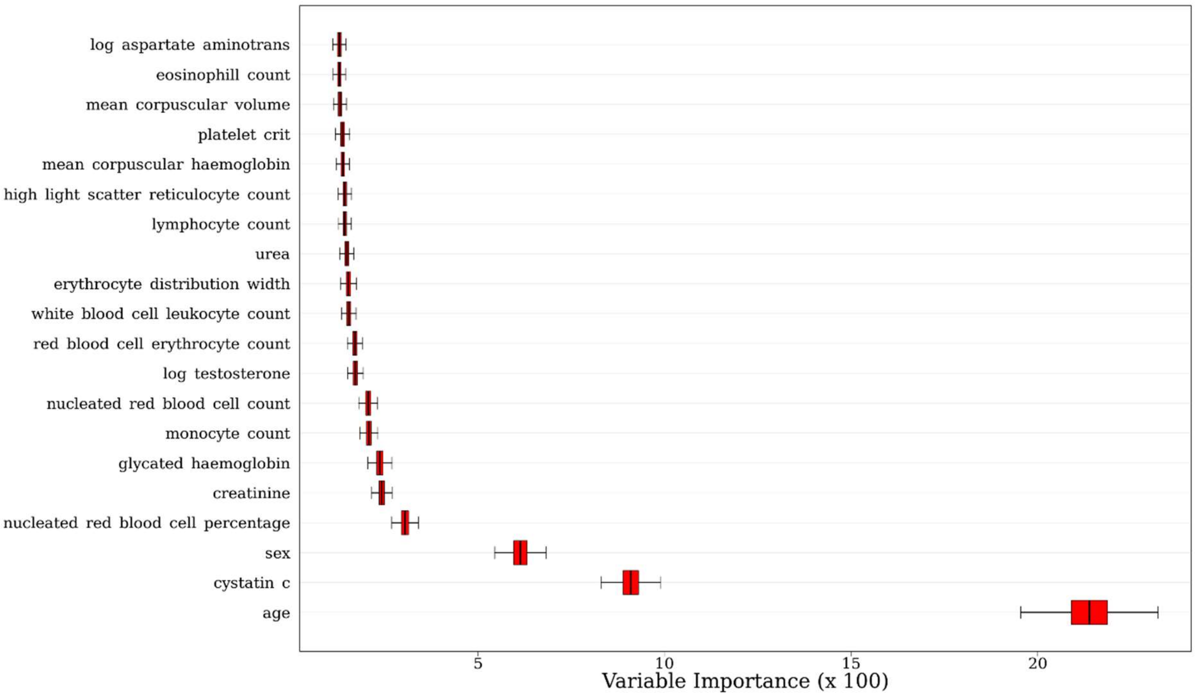


Fig SF2: Variable Importance (VIMP) values from the RSF, for the top 20 variables, in ascending order. As seen in the Full ENC model, cystatin C appears to be the biomarker of primary importance. The VIMP scores suggest that cystatin C provides a notable 0.09 [0.08 - 0.1] uplift to the RSF C-Index.

### Supplementary Note

#### Results of sex-stratified sensitivity analysis

Fewer biomarkers were stably selected in each of the sex-stratified 100-iteration Elastic-Net Cox models, compared to the Full ENC model. Selection proportion plots can be seen in Figures SN1 and SN2. Whilst 27 variables were stably selected in the full model, 22 variables were stably selected in the male-only model and 15 were stably selected in the female-only model. The standardised coefficients and confidence intervals for each of the sex-stratified models are illustrated in Fig SN3, and Table SN4 displays a side-by-side comparison of selected variables in each model and their corresponding effect sizes on the log hazard scale. The variables with the largest standardised effect sizes generally appear in both the male- and female-only models. Cystatin C once again displays the largest standardised coefficient in both models. However, it is interesting to note that (the log of) alanine aminotransferase – a variable with a large protective effect in the full model - was selected in fewer than 10 out of the 100 iterations in the female-only model (Fig SN2), suggesting that this variable is only associated with male specific mortality. Other variables, such as albumin, alkaline phosphate, and urea, also showed a strong sex-specific association with mortality. For variables which were selected by either (or both) of the stratified models, most coefficient values reflected the coefficients generated by the Full ENC model, and no major movements are identified.

Using the sex-stratified models, we produced predicted values and C-index values for each, on the test set. The results can be seen in the bottom row of Table SN4. Whilst we may expect C-index values in single-sex populations to be lower than in a mixed population, the male-only model produced a C-index higher than the Full ENC model, of 0.764 [0.754 - 0.773], whilst the female-only model produced a C-index of 0.729 [0.715-0.743]. Given that these predictions are made within single-sex groups, with sex – a major differentiator of mortality – no longer present, these results are both comparably high. Interestingly, when using the Full ENC model to make predictions for males only, and separately for females only, the respective C-indices were identical to each of those produced by the sex-stratified models, suggesting that sex-stratified models do not necessarily produce more accurate predictions than does the Full ENC model, even within same-sex group.

We compared the BAA estimates from the Full ENC model to estimates generated from the sex-stratified models. A comparison of the density plots can be seen in Fig SN5, and a scatterplot examining the correlation between the two estimates is shown in Fig SN6. The range and shape of the distributions are largely identical between the two approaches. Males appear to have a wider range of BAA values. The correlation between the BAA estimates under the two approaches is approximately 1 for males ($p < 0.0001$). However, for females, the Full ENC model produces slightly higher BAA values, on average, than the female-specific model. A possible reason for this overestimation may be the inclusion of variables which do not appear to be associated with female mortality - as indicated in Table SN4 - such as alanine aminotransferase, albumin and urea. However, the correlation between the two estimates for females is still high, at 0.94 ($p < 0.0001$).

Based on the observations made during the sensitivity analysis, we produced one final Cox model on the full training set, using the previously derived Full ENC model as the base. For this model, we removed variables which were selected by neither the male- nor female-only models. Also, for variables which showed a strong sex-specific association with mortality, we included an interaction term with sex. These variable-specific treatments are indicated in the last column of Table SN4. The C-index of this model based on predictions on the test was 0.762 [0.754-0.77] – exactly identical to the C-index achieved by the Full ENC model, suggesting that these modifications do not improve predictive performance. Similarly to the above, we also produced BAA estimates using this model, and compared them to the estimates produced by the sex-stratified models. The results were again very similar to Full ENC model: neither the male nor female correlation values improved, and BAA estimates for females were still slightly overestimated compared to the female-specific model.

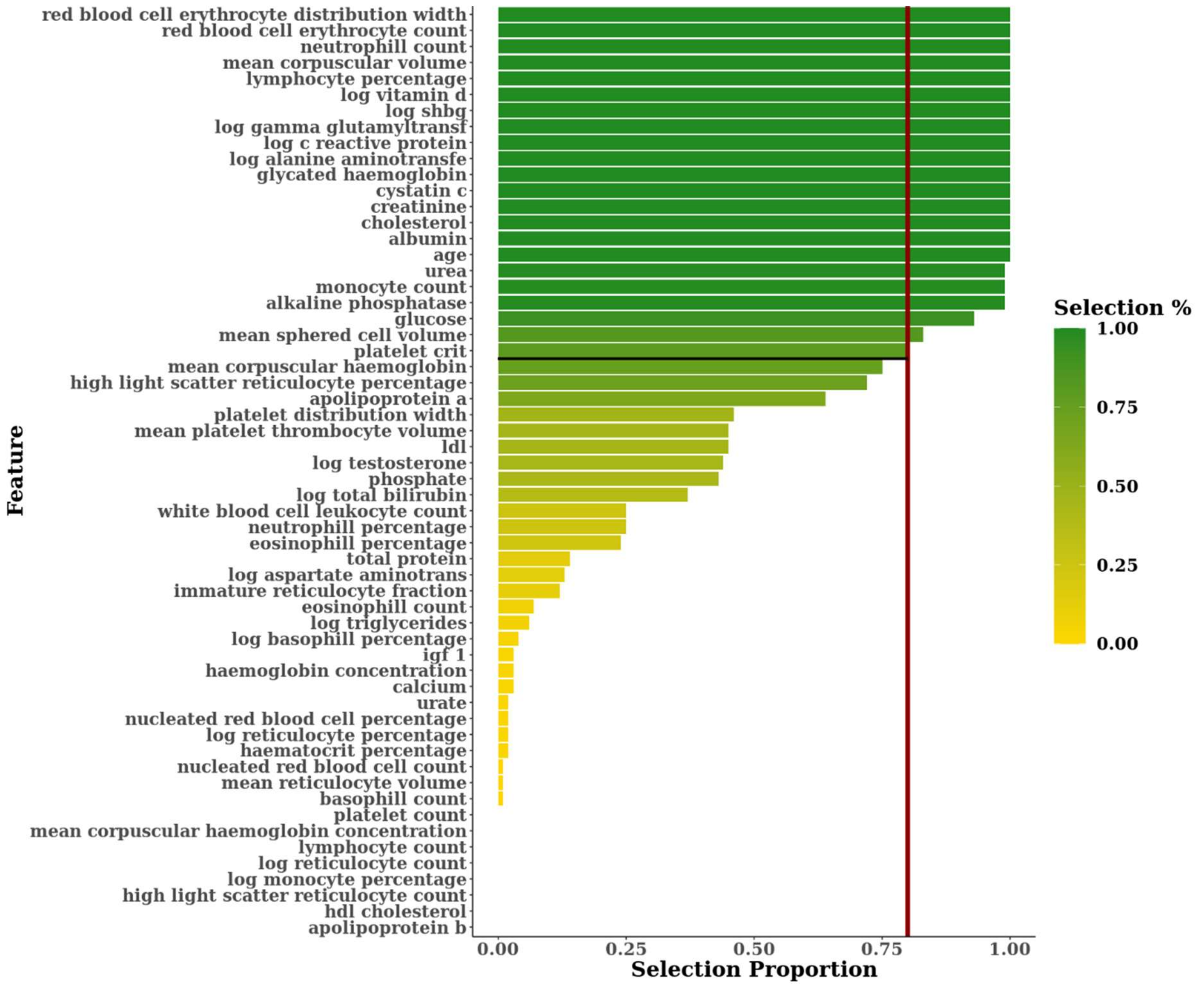


Fig SN1: Selection proportions of each feature as a percentage of the 100 Elastic Net iterations performed, ranked from highest to lowest, for the male-only model. The selection threshold of 80% is indicated in red. 22 variables were stably selected.

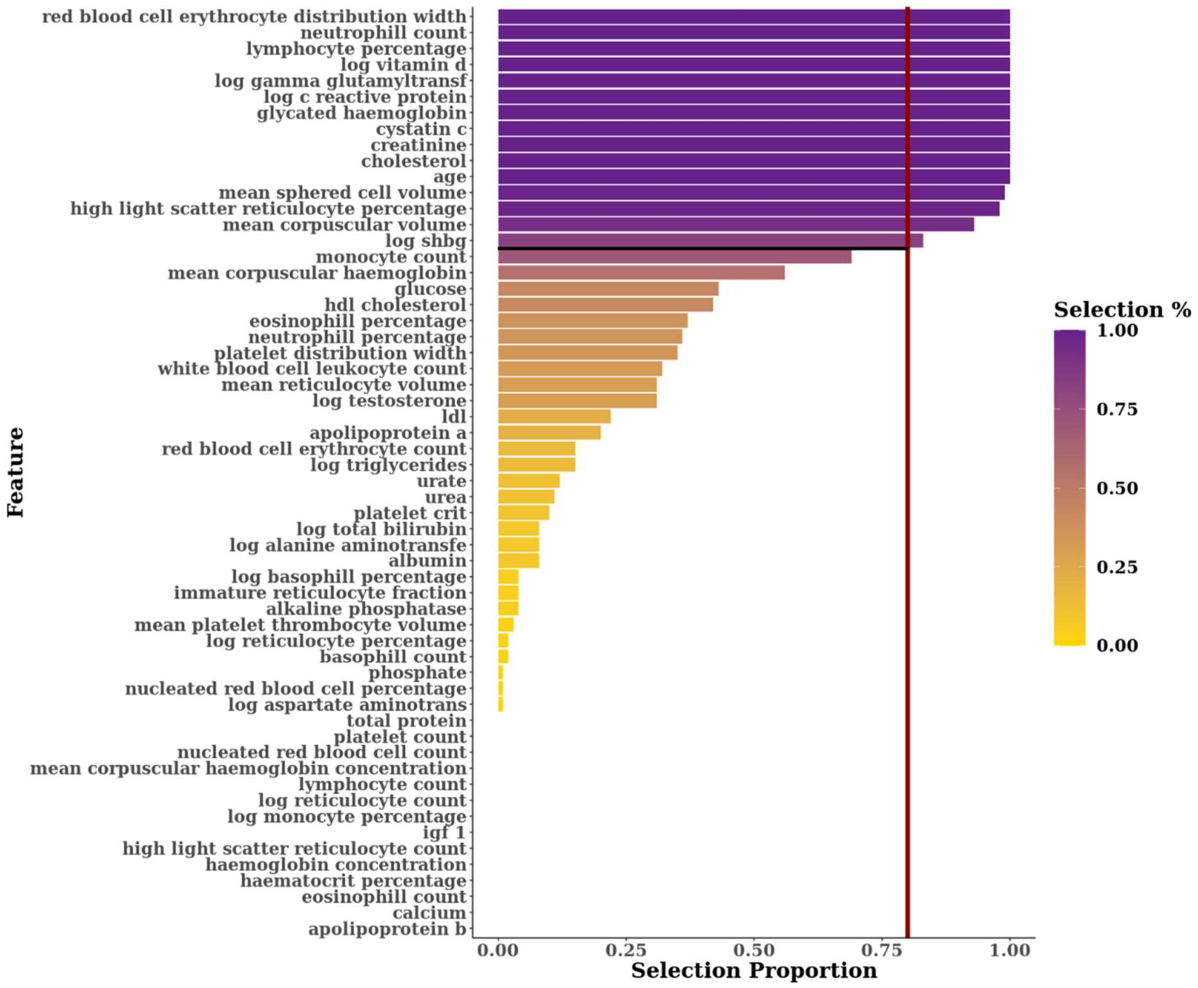


Fig SN2: Selection proportions of each feature as a percentage of the 100 Elastic-Net iterations performed, ranked from highest to lowest, for the female-only model. The selection threshold of 80% is indicated in red. 15 variables were stably selected.

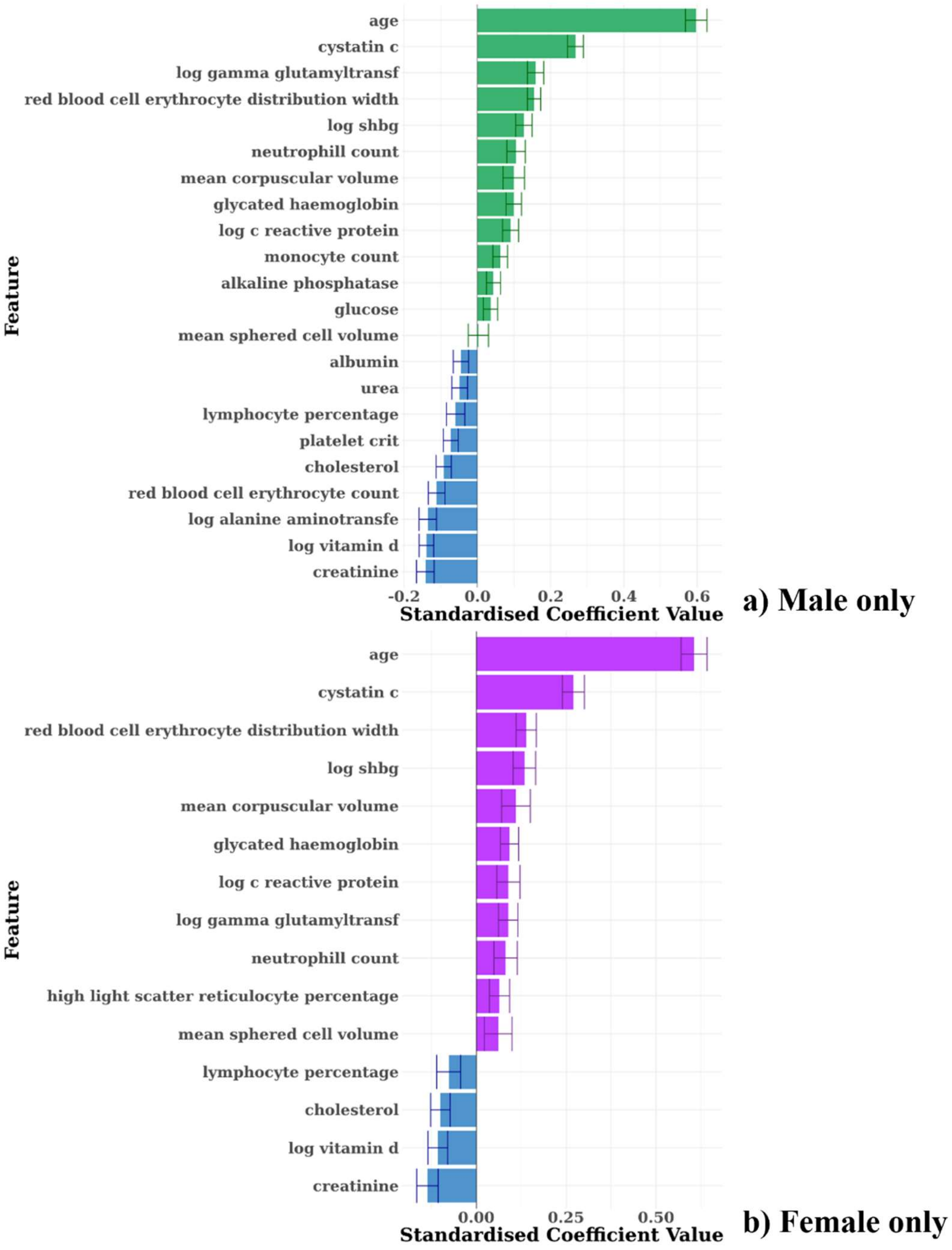


Fig SN3: The standardised coefficients of the Cox models developed using stably selected variables for (a) males and (b) females separately, ranked in descending order. Coefficients are standardised (i.e. re-scaled) by multiplying by the standard deviation of the variable concerned. 95% confidence intervals are indicated by the overlaid bars. Variables with the strongest effect sizes are reasonably consistent across both sexes. Cystatin C appears to have the strongest effect sizes for both males and females, as was the case in the Full ENC model.

Table SN4: Comparison of coefficient values between the Full ENC model trained on both sexes and the sex-specific Cox models. Colours indicate the magnitude and direction of coefficients, with red indicating that higher levels increase mortality risk, whilst blue indicates a protective effect. Grey blocks indicate the selection proportion for variables which did not breach the 80% selection threshold. A comparison of concordance values is displayed in the last row. The last column indicates the course of action taken based on this comparison, to develop a model with sex interactions and explore the impact on C-index of predictions. Measurement units per variable are displayed in Supplementary Table 1. No additional variables were selected in the stratified models which were not selected in the full model. Some variables which were stably selected in the full model, were not selected by either of the stratified models (apolipoprotein-A, mean corpuscular haemoglobin, and platelet distribution width). Albumin, alkaline phosphatase and urea were each selected by 100% of iterations of the male model, whilst being selected in fewer than 10% of female model iterations. Conversely, high light scatter reticulocyte percentage was stably selected for females only.

| | | Sensitivity Analysis | | |
|---|---|---|---|---|
| **Feature** | **Full Cox Model Coefficients** | **Male only E-NET Cox model** | **Female only E-NET Cox model** | **Subsequent Treatment** |
| Age | 0.075 | 0.073 | 0.075 | |
| Albumin | -0.009 | -0.018 | <10% | Sex interaction |
| Creatinine | -0.012 | -0.011 | -0.013 | |
| Glucose | 0.034 | 0.041 | <50% | Sex interaction |
| log CRP | 0.080 | 0.089 | 0.082 | |
| Lymphocyte % | -0.010 | -0.008 | -0.011 | |
| Mean cell vol | 0.016 | 0.024 | 0.026 | |
| Red Cell Distribution Width | 0.197 | 0.207 | 0.160 | |
| Alkaline Phosphatase | 0.001 | 0.002 | <10% | Sex interaction |
| White Blood Cell Count | <50% | <50% | <50% | |
| Apolipoprotein A | -0.198 | <70% | <30% | Remove variable |
| Cholesterol | -0.078 | -0.090 | -0.104 | |
| Cystatin-C | 1.877 | 1.917 | 1.926 | |
| Glycated Haemoglobin | 0.019 | 0.018 | 0.020 | |
| High Light Scatter Reticulocyte % | 0.178 | <80% | 0.324 | |
| log Vitamin D | -0.260 | -0.296 | -0.229 | |
| log Alanine Aminotransferase | -0.300 | -0.327 | <10% | Sex interaction |
| log Gamma-Glutamyltransferase | 0.264 | 0.278 | 0.160 | |
| log Sex Hormone Binding Globulin | 0.310 | 0.308 | 0.278 | |
| Mean Corpuscular Haemoglobin | 0.026 | <80% | <70% | Remove variable |
| Mean Sphered Cell Volume | 0.007 | 0.001 | 0.012 | |
| Monocyte Count | 0.340 | 0.364 | <80% | |
| Neutrophill Count | 0.070 | 0.078 | 0.061 | |
| Platelet Crit | -1.356 | -1.709 | <20% | Sex interaction |
| Platelet Distribution Width | 0.060 | <50% | <50% | Remove variable |
| Red Blood Cell Count | -0.215 | -0.309 | <20% | Sex interaction |
| Sex (male) | 0.590 | NA | NA | |
| Urea | -0.030 | -0.037 | <20% | Sex interaction |
| **C-Index on Test Set [And 95% CI]** | All:<br>0.762 [0.754 – 0.770]<br>Males only:<br>0.764 [0.755 - 0.773]<br>Females only:<br>0.729 [0.716 - 0.743] | 0.764 [0.754-0.773] | 0.729 [0.715-0.743] | 0.762 [0.754-0.770] |

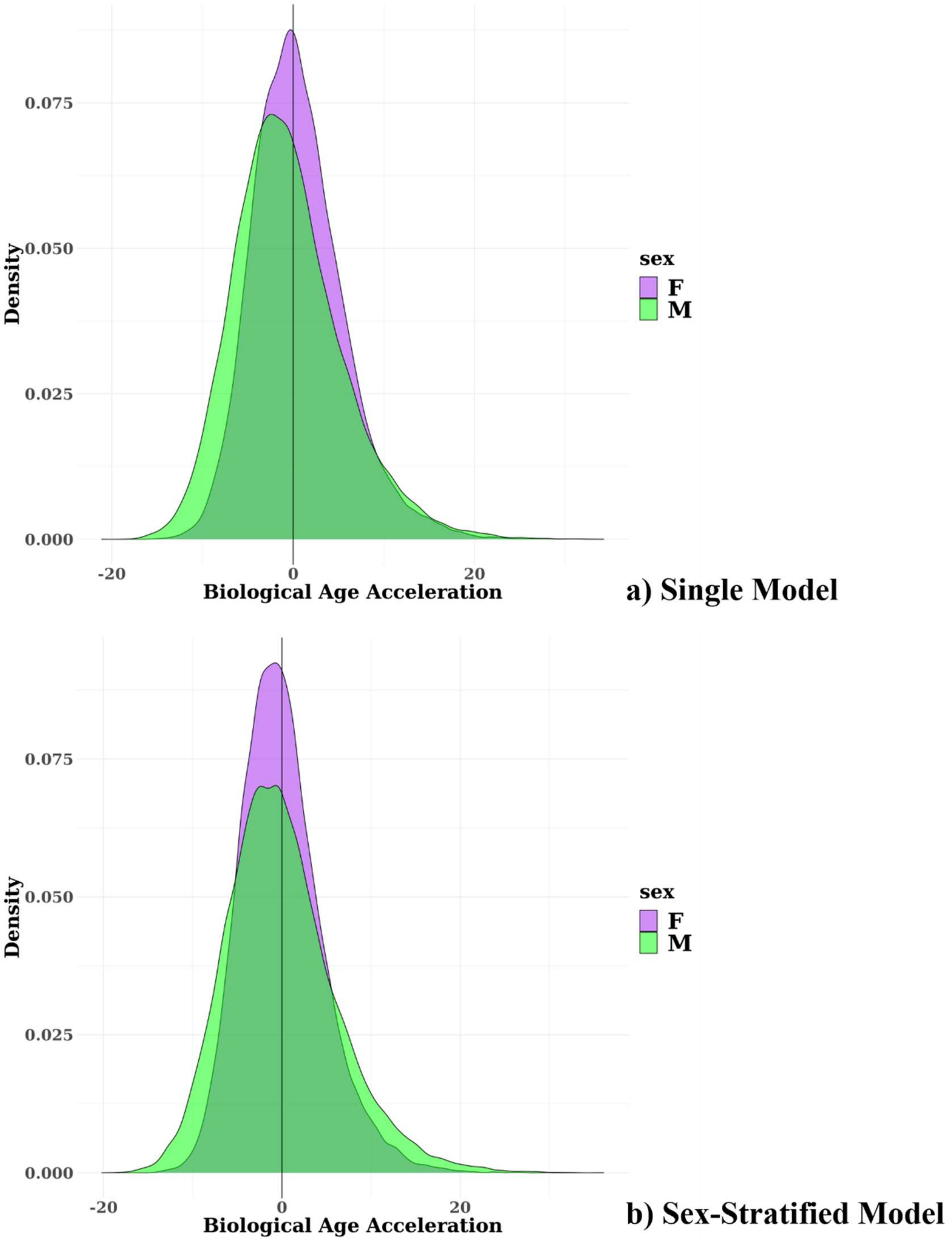


Fig SN5: Distributions of BAA by sex based on the single, Full ENC model (a) compared to BAA estimates generated by sex-specific models (b). The range and shape of the distributions are largely identical between the two approaches. Males appear to have a wider range of BAA values.

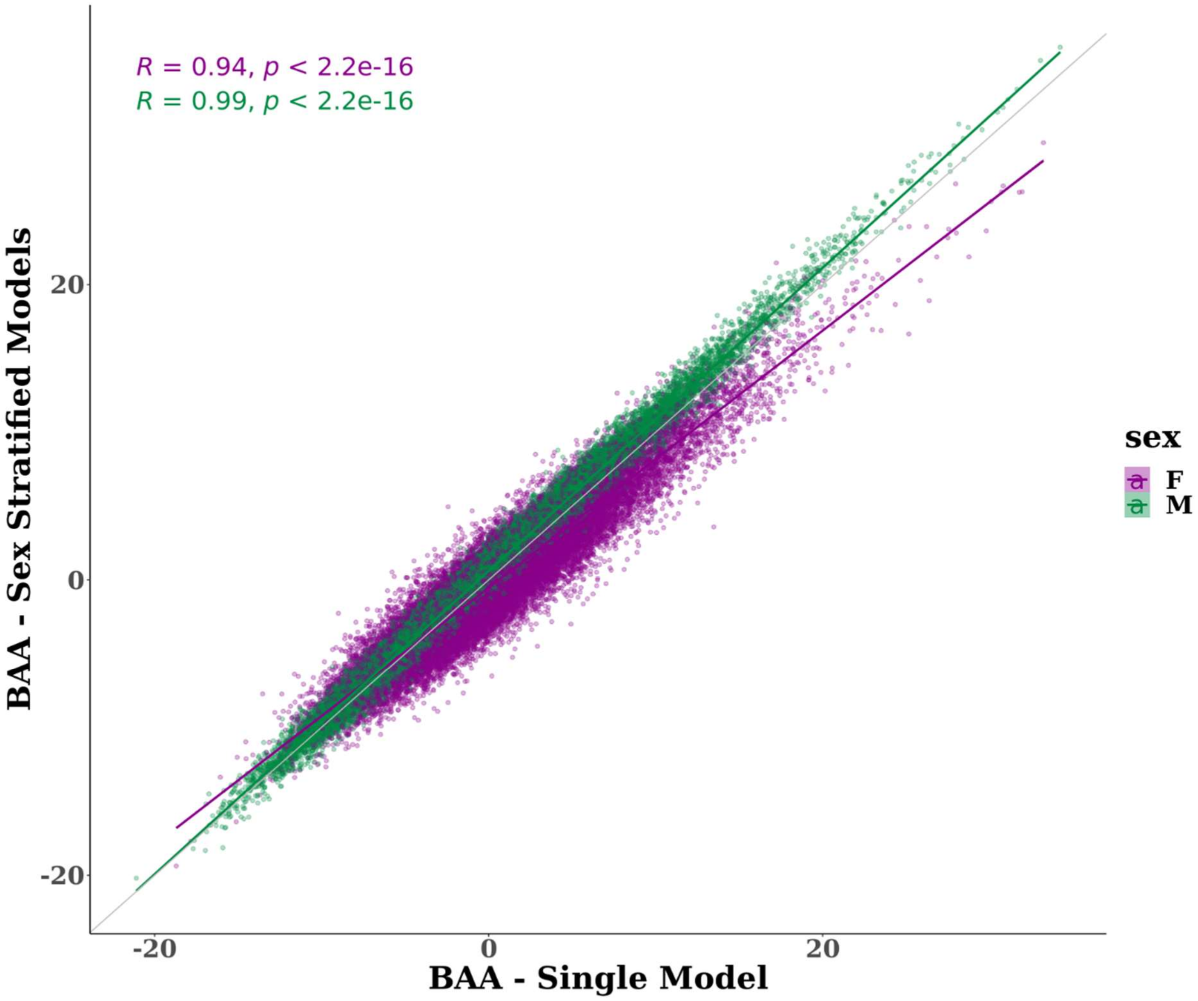


Fig SN6: Scatterplot indicating the correlation between BAA estimated using the single Full ENC model vs sex-specific models. BAA for males was almost identical across approaches, whilst the single model may overstate BAA estimates for females.
